# Owning the narrative: Exploring the impact of a creative storytelling intervention on Stigma and Empowerment among persons affected by Leprosy in Pakistan

**DOI:** 10.64898/2026.02.23.26346453

**Authors:** Nervana Ibrahim, Anil Fastenau, Abdul Salam, Fabian Schlumberger, Matthew Willis, Lucy McKane, Ali Murtaza, Maaike Seekles, Laura Dean, India Hotopf

## Abstract

**Introduction:** Despite being curable, leprosy-related stigma in Pakistan persists, undermining dignity, delaying care, and hindering progress toward zero-leprosy targets. Empowerment is critical in counteracting stigma and restoring agency among affected persons. Participatory, contact-based storytelling has potential to reduce stigma and strengthen empowerment, but evidence remains limited. This study evaluates a participatory storytelling intervention involving persons affected by leprosy in Karachi.*Objectives* This study investigates how a participatory storytelling intervention influences stigma and empowerment among persons affected by leprosy in Pakistan. It explores changes in participants’ experiences, examines the mechanisms through which storytelling engages with stigma and identifies practical insights to strengthen future intervention design.

**Methods:** We conducted a qualitative study following the 2024–25 participatory storytelling pilot at MALC, Karachi. Persons affected by leprosy who participated in the intervention were purposively sampled (age 30-65 years); sixteen participated in photovoice and fourteen completed in-depth interviews (IDIs) (thirteen of which had also participated in photovoice) (total n= 17). Data were analysed thematically using the Social Ecological Model (SEM).

**Results:** The programme supported a shift from shame and concealment to greater self-acceptance, confidence and openness. Peer groups and creative storytelling built solidarity, a sense of belonging and transferable skills, while organisational endorsement enhanced participants’ visibility and roles. However, stigma remained reinforced by community misconceptions, gender norms and practical barriers. Overall, the intervention’s greatest effect was at the individual level, reducing self and anticipated stigma, with intrapersonal empowerment emerging as the primary gain.

**Conclusion:** Participatory storytelling enhanced identity and empowerment, but broader social change remained constrained by gender norms, visible disability, and structural barriers.

Strengthening impact requires sustained community contact, supported peer educators, inclusive outreach to women and persons with visible impairments, and links to socioeconomic opportunities. Embedding peer-led storytelling within skin NTD services and national and WHO strategies can support community ownership and sustainability.

**Author summary:** Leprosy is a curable disease, yet many people affected by it continue to experience stigma that affects their mental health, confidence, relationships and health seeking behaviour. In Pakistan, stigma remains a major barrier to achieving zero-leprosy goals and improving quality of life. Our study explored the degree to which a participatory storytelling intervention, where persons affected by leprosy share their experiences through creative methods, supported empowerment and reduced stigma.

We conducted a qualitative study with participants involved in a storytelling pilot intervention in Karachi, using interviews and photovoice to explore their experiences. Participants described the transition from shame and secrecy toward openness, greater self-confidence and self-acceptance. The storytelling process encouraged peer support, strengthened social connections and helped participants develop new skills and a stronger sense of identity. However, persistent community misconceptions, gender norms, visible disability and economic constraints limited broader social change.

Our findings suggest that participatory storytelling interventions present a promising approach for reducing internalised stigma and strengthening individual empowerment among persons affected by leprosy. To achieve wider impact, storytelling initiatives should be combined with sustained community engagement, inclusive outreach and integration into skin neglected tropical disease (NTD) services and broader social support programmes.

## 1. Introduction

Leprosy, a chronic infectious condition caused by *Mycobacterium leprae*, primarily affecting the skin and peripheral nerves (1). Despite major reductions in global incidence since the 1980s, there were 172, 717 new cases reported in 2024 (1, 2). As a Neglected Tropical Disease (NTD), leprosy remains underfunded and under-researched and disproportionately affects marginalised communities living in poverty (3, 4). Like many skin NTDs, leprosy causes social stigma and exclusion, contributing to poor mental health, reduced employment opportunities and long-term psychosocial harm (5) (6, 7).

While medical management of leprosy has advanced, psychosocial dimensions remain inadequately addressed (6, 8, 9). Stigma continues to undermine public health efforts and threatens the global goal of achieving zero leprosy (3, 10). Global efforts, including the World Health Organisation (WHO)’s Global Leprosy Strategy 2021–2030 ‘Towards Zero Leprosy’, aim not only to reduce disease burden through early detection and treatment, but also to tackle stigma, discrimination and social exclusion (11). These priorities support the Sustainable Development Goals (SDGs) by addressing social determinants of stigma and advancing equity for vulnerable populations affected by NTDs (12). Despite this, there is a dearth of evidence on which interventions are most effective at reducing stigma and addressing psychosocial needs (8, 13–15).

Health-related stigma refers to the negative social judgements and devaluation of individuals based on a particular health condition (7). Building on Goffman’s definition, stigma is understood as an attribute that discredits individuals and can lead to exclusion (16). In leprosy, stigma is reinforced by visible deformities and historical associations with isolation (17, 18). Misconceptions persist despite effective treatment, fuelling concealment and discouraging care-seeking, which results in delayed diagnosis, greater disability and escalating psychosocial harm (15, 17, 19, 20). Stigma does not operate solely as an individual psychosocial burden: it is embedded in structural inequities that shape access to information, services and social protection. Gender, disability, socioeconomic status and wider social marginalisation can intensify exposure to stigma and constrain timely care seeking and care access, with consequences for disability and exclusion (21, 22).

Empowerment is a central concept for challenging stigma and promoting resilience. Defined as comprising intrapersonal, interactional and behavioural components, empowerment strengthens autonomy, self-efficacy and the capacity to influence one’s circumstances (23). For persons affected by skin NTDs such as leprosy, empowerment can be both an outcome and a mechanism through which individuals reclaim dignity, strengthen agency and counteract stigma (21). However, empowerment remains a neglected domain within leprosy control (24).

Efforts to reduce stigma have included information, education and communication (IEC), community-based support and narrative or participatory approaches (8). Evidence suggests multi-component interventions generally yield better outcomes than education alone (15, 25, 26). Storytelling and participatory methods have gained attention for their potential to transform both self-perception and public attitudes (9, 27). Narrative strategies integrate factual content with emotional and experiential components, which can reduce resistance and foster attitude and behaviour change (28–30). Participatory tools such as drama, photovoice and video can position persons affected as active contributors rather than passive subjects, supporting narrative ownership and authenticity (31–36). Despite growing interest, rigorous evaluation of storytelling interventions in leprosy remains limited, with much evidence stemming from small-scale or pilot studies and limited long-term follow-up (9). Research in Pakistan remains scarce, with no published evaluations of leprosy-related stigma interventions (37).

Pakistan achieved elimination of leprosy as a public health problem in 1996 and is classified as low-endemic (38). Despite this progress, 20.7% of newly diagnosed patients present with Grade 2 Disability (G2D), exceeding the global average of 5.5% and underscoring persistent delays in diagnosis across the country (39, 40). Stigma and misconceptions are among key drivers of these delays, increasing the risk of disability and continued transmission (20, 37). In response, Pakistan has developed a Zero Leprosy Roadmap aimed at eliminating leprosy by 2030, prioritising not only zero disease and disability but also zero stigma, including the meaningful inclusion of affected persons in campaigns to challenge myths and promote social acceptance (41).

Within this context, a participatory stigma-reduction intervention was piloted in Karachi in 2024, incorporating participatory storytelling and contact-based activities alongside health education, with engagement of persons affected by leprosy from communities across Sindh (42). As next steps are considered, it is essential to assess the intervention’s effectiveness and impact. This study addresses the current evidence gap in Pakistan by evaluating this participatory storytelling intervention. It explores how the intervention influences stigma and empowerment among persons affected by leprosy and examines the processes through which storytelling may contribute to stigma reduction and empowerment. The study was guided by the research question: How does a participatory storytelling intervention influence stigma and empowerment among persons affected by leprosy in Pakistan? It further aims to identify key enablers and barriers and to generate evidence-based recommendations to strengthen intervention design and implementation.

By focusing on both perceived impacts and mechanisms, this research contributes practice-relevant evidence for leprosy programmes seeking to move beyond IEC-only approaches toward more participatory, person-centred strategies. It explicitly situates stigma within wider inequities affecting access to care and social inclusion and considers how intersecting factors such as gender, disability and socioeconomic marginalisation may shape experiences of stigma and empowerment. Findings are intended to inform refinement and potential scale-up of the intervention and to support national and global commitments to zero leprosy, including the SDG agenda’s emphasis on equity and leaving no one behind.

## 2. Methods Setting

In 2024, a one-year pilot programme to address leprosy-related stigma was launched in Karachi, Pakistan, through a collaboration between the ILEP, GLRA and the Marie Adelaide Leprosy Centre (MALC). The intervention was implemented at MALC and integrated participatory storytelling with community-based group sessions delivered through a peer support group (PSG), combining skills-building and creative sessions with health education and advocacy (e.g., disability rights and self-care). It incorporated contact-based components that brought persons affected into direct interaction with key stakeholders, including MALC staff and selected community members. Public engagement activities also included awareness sessions with participants’ stories shared through creative performances and a filmed component documenting their lived experience.

The lead author (NI) is a medically trained doctor (MBBCh, MPH) who conducted this study as part of her Masters in Public Health programme and completed extensive training in qualitative research methods. The co-researchers all have extensive experience and training in qualitative research and skin NTDs.

This study adhered to the Consolidated Reporting Criteria For Qualitative Studies (COREQ) (S1 Appendix).

### Study design

This qualitative naturalistic study used inlZldepth interviews (IDIs) and photovoice to explore experiences of persons affected (S2 and S3 Appendices). Photovoice provided additional participant engagement and offered visual reflection on their experiences (43). It is valuable as a participatory research method because it enables persons affected to challenge dominant narratives and support empowerment through collective reflection, while functioning not just as a data collection method but as a process that centres them as experts and makes psychosocial experiences visible to health systems (44) (45). An intersectional lens was applied throughout the research cycle, to explore how multiple intersecting social identities, including gender and disability, influenced participants’ experiences (46, 47). In line with the naturalistic paradigm, the study embedded inductive analysis and purposive sampling (48).

The study drew on an integrated theoretical framework incorporating stigma and empowerment perspectives to inform the topics guides and initial analytic sensitising concepts (i.e., anticipated domains of stigma mechanisms and empowerment processes) before situating findings within the SEM (7, 23, 49, 50). The SEM informed interpretation by conceptualising influence across five levels – individual, interpersonal, community, institutional and structural – supporting analysis beyond individual-level change to the wider context in which stigma and access to care are produced (50–52) (Figure 1).

**Figure 1.**
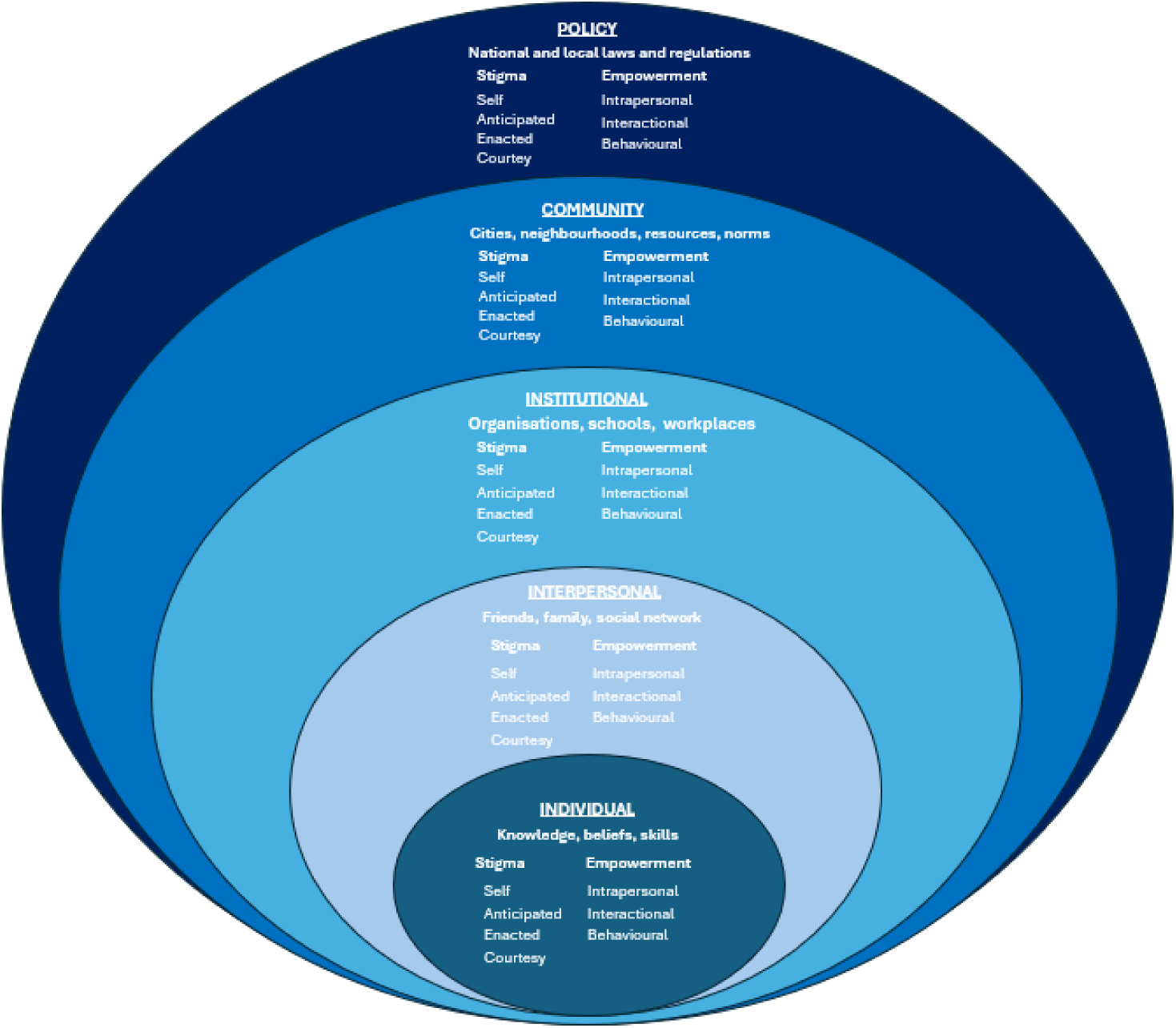
Analytical framework: empowerment and stigma across the levels of the social ecological model (Source: authors own)

### Recruitment and sampling

Recruitment occurred between 30 May–20 June 2025, with data collection spanning a 4-week period in June 2025. Participants were purposively sampled based on being affected by leprosy, participating in the intervention and being 18 years or older (Table 1). Sixteen participated in photovoice and 14 completed IDIs (13 of which had also participated in photovoice). Applying maximum variation sampling, captured a range of experiences and variation was sought across gender and age, and practically extended to disability status or visible impairments, length of time since diagnosis, and differences in social circumstances (e.g. household roles and livelihood context), using programme records and staff knowledge to identify eligible participants with differing profiles.

**Table 1:**
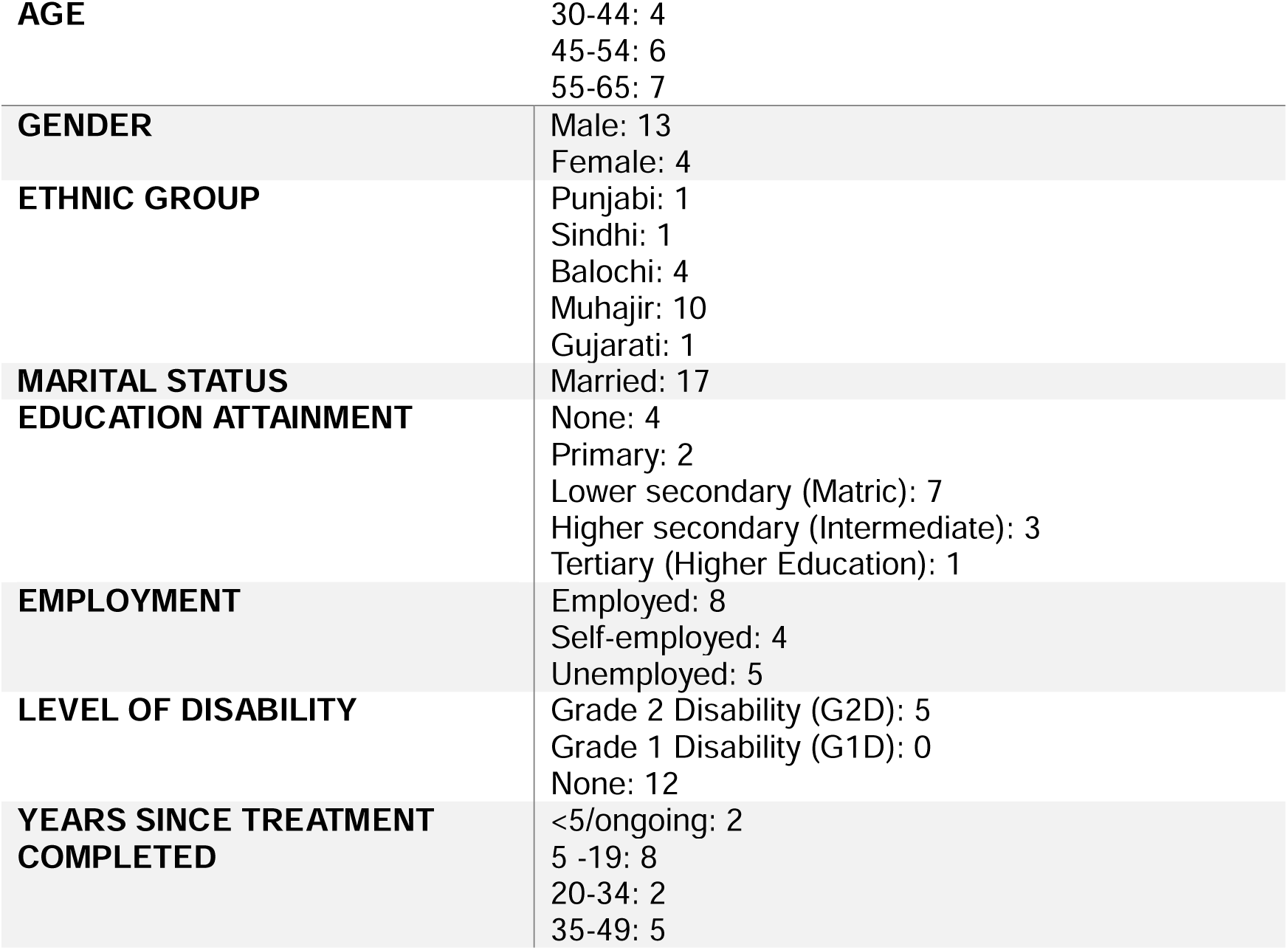
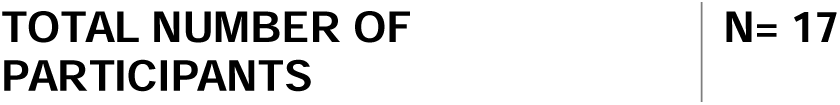
Participant demographics.

|  |  |
| --- | --- |
| **AGE** | 30-44: 4<br>45-54: 6<br>55-65: 7 |
| **GENDER** | Male: 13<br>Female: 4 |
| **ETHNIC GROUP** | Punjabi: 1<br>Sindhi: 1<br>Balochi: 4<br>Muhajir: 10<br>Gujarati: 1 |
| **MARITAL STATUS** | Married: 17 |
| **EDUCATION ATTAINMENT** | None: 4<br>Primary: 2<br>Lower secondary (Matric): 7<br>Higher secondary (Intermediate): 3<br>Tertiary (Higher Education): 1 |
| **EMPLOYMENT** | Employed: 8<br>Self-employed: 4<br>Unemployed: 5 |
| **LEVEL OF DISABILITY** | Grade 2 Disability (G2D): 5<br>Grade 1 Disability (G1D): 0<br>None: 12 |
| **YEARS SINCE TREATMENT COMPLETED** | <5/ongoing: 2<br>5 -19: 8<br>20-34: 2<br>35-49: 5 |

Recruitment was facilitated by the MALC field officer, who provided PSG members with PIS forms and introduced them to the study. The lead researcher (NI) attended a subsequent PSG to meet prospective participants and answer queries. Afterwards, the co-ordinator followed up with participants to collect informed consent. A few eligible participants were unable to take part due to time constraints or other personal commitments.

### IDI methodology

#### Development and validation of topic guide

The interview topic guide covered themes such as personal narratives and identity; stigma and perceptions, empowerment and agency; and reflections on the intervention process itself (see S2 Appendix for IDI topic guide). It was developed based on the research objectives and relevant literature, reviewed and piloted with LSTM colleagues for clarity and contextual relevance, and validated by senior colleagues from both LTSM and MALC. The topic guide was also iteratively refined during data collection to incorporate emerging insights.

#### Data collection

IDIs enabled in-depth exploration of participants’ experiences and perceptions and supported open expression in a flexible conversational format (53). Interviews were conducted in person on the MALC premise between 2 -25 June 2025. NI conducted interviews with live consecutive interpretation in Urdu by trained translators. Three translators supported data collection; all had experience in qualitative interviewing, and two also held implementation roles within the intervention (local programme co-ordinator; international programme co-ordinator). Interviews lasted 60-90 minutes, were audio recorded with consent and complemented by field notes.

#### Photovoice methodology

Participants attended an initial training session on the photovoice methodology, including ethical photography, consent, confidentiality and practical camera or phone use (54) (see S3 appendix for the topic guide). They were invited to take photographs illustrating how the intervention influenced experiences of stigma and empowerment, guided by broad themes aligned with the interviews (identity, stigma, empowerment and reflections on storytelling). Participants typically took up to ten photographs over approximately three weeks, with brief check-ins to support participation and address practical or ethical issues.

Prior to the group discussion, NI met participants individually to review images, support caption development (using the SHOWeD) method and help select two to three photographs, that participants felt comfortable sharing. In facilitated group discussions, participants presented their selected images and explained their meaning, followed by wider discussions on how the challenges highlighted could be addressed. Prompts adapted from the SHOWeD framework were used flexibly to support reflection. The group discussion also entailed co-analysis, wherein participants were supported to collaboratively cluster photographs into thematic areas. Group discussions were audio recorded, with field notes and observations captured. Photovoice data was analysed thematically and triangulated with IDI findings to contextualise themes.

An internal exhibition of selected photographs and narratives was held at MALC on 27^th^ June 2025 for MALC staff and organisational leadership, providing participants with a platform to share perspectives and enabling stakeholders to engage with visual accounts of stigma, empowerment and change.

#### Data analysis

Data analysis was conducted by NI, with supervision and support from IH. Audio recordings were transcribed verbatim by NI based on the English interpretation provided during the interviews. Transcripts were quality-checked through repeated listening and meaning checks with translators. Data were analysed using the thematic analysis (55), supported by NVivo 14.

Coding combined deductive and inductive approaches. A preliminary codebook was developed deductively from the topic guide and the analytic frameworks (stigma, empowerment and SEM domains), then iteratively expanded with inductive coding arising from the data. NI conducted primary coding across the dataset. To strengthen analytic rigor, a senior member of the research team reviewed the developing codebook and a subset of coded transcripts; any ambiguities or disagreements were discussed and resolved through consensus, with refinements documented in the codebook (S4 Appendix). Themes were developed through iterative comparison across participants and methods, refined through team discussion, and interpreted in alignment with the theoretical framework (Figure 1) . An intersectional lens was applied throughout, with close attention paid to how identity factors such as gender, shaped experiences. Photovoice data (images, captions and group discussion transcripts) were analysed alongside IDIs to triangulate and contextualise themes. Key themes were also co-analysed during the photovoice reflective group discussions through interactive discussion between NI and participants.

#### Ethics

Ethical approval was obtained from the Liverpool School of Tropical Medicine (ID: MSc25(07)) and MALC Ethics Review Committee (ID: MAC/ERC//03/2025). Informed consent was obtained for participation, audio recording and photography as well as sharing of photos. Participation in the study was voluntary, and it was emphasised that taking part would not affect the services respondents received from MALC. The purpose of the research was clearly explained, including the objectives, processes and participants’ rights (including the right to withdraw), with time provided for questions and clarification. Written and verbal consent were obtained, with the voluntary nature of participation emphasised. For non-literate participants, consent was documented using a thumbprint as a signature (see S5 and S6 Appendices). The consent process was carried out with translators present throughout the process to ensure participants fully understood the information provided. Considering the stigmatising nature of skin NTDs, NI worked alongside local staff and was trained in recognising and addressing signs of distress that persons affected may have due to discussing sensitive issues. Participants were debriefed afterwards to ensure their well-being and to provide an opportunity to discuss any concerns. Counselling support services were available to participants if needed. All data were anonymised, stored securely using password-protected storage and handled confidentially.

#### Positionality and reflexivity

NI is a medically trained female from a similar cultural background and was thus able to build rapport and contextual understanding. However, as an outsider there was reduced preconceptions, which facilitated greater objectivity. The translators’ dual role as programme implementers and interpreters was acknowledged as a potential bias and mitigated through reflexivity and participant checking.

Additionally, participants were reassured that they could speak freely without repercussions.

## 3. Results

This section presents qualitative findings of the leprosy storytelling intervention, interpreted through an integrated theoretical framework incorporating stigma and empowerment perspectives, and situated within the SEM to explore:(1) its impact on stigma and empowerment (2) factors influencing its effectiveness and (3) participant recommendations for improvement.

### 3.1 Impact on Stigma and Empowerment

#### Living with Leprosy: Life before the Intervention

Before joining the programme, participants described prolonged and uncertain illness trajectories, misdiagnosis and financial strain, often compounded by loss of work and educational opportunities. Community-level enacted stigma included avoidance, verbal abuse and exclusion from work or public transport, with men with visible Grade 2 Disability (G2D) particularly vulnerable to public stigma.

Stigma within immediate families was generally low. However, one woman was temporarily evicted due to contagion fears and reaccepted after targeted health education. For some men, female relatives buffered enacted stigma, highlighting the gendered role of women in providing acceptance and care.

Anticipated stigma shaped everyday choices, with concealment and non-disclosure adopted as protective strategies, especially when deformities or treatment effects were visible. One woman emphasised fear of disclosure for married or younger women, who hide their condition from husbands or in-laws due to anticipated stigma. Those with G2D reported heightened enacted stigma, facing persistent questioning and scrutiny. Self-stigma manifested as sadness, fear and shame, even escalating to suicidal ideation for one young male. Health-system encounters sometimes compounded stigma through misdiagnosis or physical distancing, delaying treatment and increasing disability risk.

Prior to the intervention, empowerment was generally limited. Participants frequently reported low confidence and negative self-perceptions. Condition-specific knowledge and service navigation skills, key aspects of interactional empowerment, were also limited and usually acquired informally through peers or family.

Photovoice participants commonly used imagery of death and lifelessness (graveyards, dry plants) to depict how they felt when diagnosed with leprosy (Figure 2). These visuals echoed the interview findings, capturing the deep sense of grief and loss of self that accompanied diagnosis.

**Figure 2.**
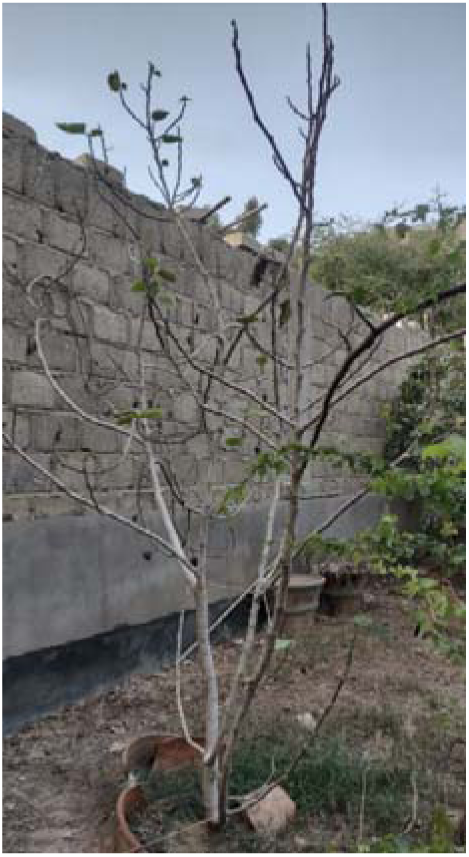
“Before the disease I was like a plant with its fruit, very productive and had happiness. When I got sick, I became like a tree that had lost it is leaves, a dry tree without any fruit”.

#### Living with Leprosy: Life after the Intervention

Findings are presented using the SEM, to explore the effect of the intervention. At each level, the impact on stigma and empowerment is examined together (S7 Appendix), since stigma reduction and enhanced empowerment exist in a symbiotic relationship, mutually reinforcing and sustaining each other.

### 3.2 Identity Transformation: From Shame to Strength

#### Reduced internal and anticipated stigma

The most significant impacts occurred at the individual level. Participants commonly described a shift from shame and concealment to self-acceptance, openness and pride, which they linked to reduced fear of judgement and increased willingness to disclose, also echoed in the photovoice discussions. Here, participants used images to describe feeling lighter, less afraid and more confident after the intervention. They described a similar emotional release, likening the training to a fire extinguisher that put out the fear and stigma burning inside them:

> *“I was feeling very light in my heart because I have already shared everything. It was like, Jwalamukhi [Volcano]. When this volcano burst out […] I have no fear, no shame, not feeling any guilt. I’m feeling like, if someone accept his crime in front of someone. I accept all those crimes, I feel lighter and so relaxed.” (P06, 40’s, Male, Government worker)*

These quotes not only illustrate the change, but also the creative analogies used to articulate their thoughts and emotions. Their use of vivid language reflects a deeper transformation, one that is evident not just in what they say, but in how they choose to express it. Several participants explicitly reported reduced anxiety (less fear, hesitation and isolation) and decreased depression, with more feelings of happiness and hope, after the intervention.

#### Enhanced intrapersonal and behavioural empowerment

Confidence emerged as the most common change, reported by all participants. They articulated a clearer sense of self, increased self-respect and a disassociation from the ‘patient’ identity. The shift in how they perceived themselves was frequently framed as moving from a diminished state to one of realised self-worth. Self-efficacy also increased, with participants expressing a renewed sense of purpose and a responsibility to support others affected by leprosy. Some reflected on how their condition, once seen as a burden, had become a source of strength. Photovoice imagery vividly reflected this transformation, with many participants framing their change in terms of courage (*Himat* in Urdu, as written in the picture) and boldness (Figure 3).

**Figure 3.**
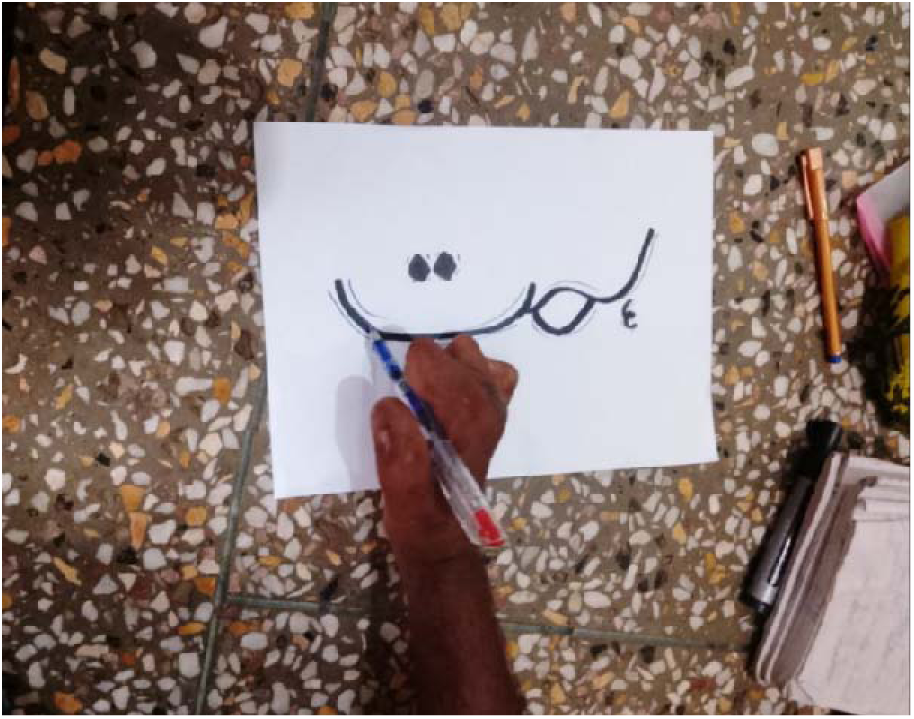
Courage, strength and determination: “There was a time, I was not able to continue with my education or even hold a pen, but after the interventions and support from MALC, I completed my education. Now even with my visibly disabled hand, with courage and determination I am now able to write and I can hold my pen. There is a lot of power in the pen. I w s taking my life forward. I was not looking at my past. With courage you can achieve anything”.

#### Knowledge and Skills Gained: Communication and Health Literacy

Participants gained concrete knowledge and transferable skills, including disability prevention and self-care, with an older male noting the importance of timely HSB to prevent worsening illness. Sharing personal journeys also exposed them to diverse experiences, encouraging reflection and deepening their understanding of leprosy and recovery.

Communication emerged as particularly transformative for most participants. They frequently described learning to listen actively, maintain eye contact and convey clear, well-structured messages tailored to their audience. Many highlighted the use of creative expression through storytelling to engage listeners more effectively.

For a couple of women, these skills extended beyond the intervention and into professional roles, particularly in community health promotion. Sharing her experience helped her connect authentically, build trust with patients and motivate vaccine uptake:

> *“My communication skills are better. I can relate to different stories, and I can convince people better[…] Before I was not listening so carefully when people were telling, and I learned how good it feels to be listened to.” (P07, 50’s, Female, Health Worker)*

##### Interpersonal Outcomes in Peer and Family Relationships

Within the PSG, participants could discuss stigma and challenges, they faced. Many shared openly without fear of judgement, laying the groundwork for trust and mutual recognition. As members affirmed each other’s struggles and celebrated small achievements, disclosure evolved into solidarity, with relationships marked by equality and respect. This transformed individual experiences into shared meaning and collective resilience.

> *“There are a lot of people they were listening to me, so nicely, so carefully and with interest. When we were all together, we are doing these activities in a group. It was very good feeling for me that if we are together and unite, so we can do a better work.”* (P12, 60’s, Male, Self-employed)

This was captured in a photovoice discussion where one participant reflected *that “every face is different, but the colour of blood is the same. The pictures are different, but the story is the same,”* highlighting how diverse personal journeys were recognised as part of a shared experience. Across IDIs and photovoice there is a consistent picture of the PSG as a space of mutual support, shared experience and collective strength.

A few reported improved family relationships, including praise and recognition following the public film screening. A male with G2D described feeling an elevated status at home, especially among in-laws who had kept their distance. For many, familial acceptance began following treatment and health education, prior to the intervention.

#### Institutional impact: Redefining Roles and Relationships

The storytelling intervention expanded the organisation’s role, redefining its relationship with persons affected and embedding empowerment institutionally. Following structured training participants engaged with MALC not as patients but as trained storytellers and peer educators. In the first public-facing stage, they shared their stories and creative pieces in public engagement events. Staff interactions reinforced this transition, by treating participants as equals. MALC’s role expanded beyond healthcare provision to advocacy aimed at reducing leprosy-related stigma. This allowed participants to contribute to public discourse, representing their experiences as active agents of change, rather than passive recipients of care.

#### Community Level: From Personal Empowerment to Community Action

At the community level, the most notable shifts occurred through contact events, informal case-finding and health education, which brought participants into direct contact with wider audiences. However, contact was mainly episodic (MALC-based events) and refugee-camp visits were one-off, undertaken by a small number of vocal peers of both genders.

Informal practices emerged, e.g. identifying suspected cases in neighbourhoods and discussing leprosy in daily conversations, illustrating the active role of persons affected in raising awareness. Reduction in enacted stigma was reported in selective public settings, such as their local neighbourhoods. A couple of men noted feeling more listened to and respected in their communities post-training:

> *“Now they listen more to us, and they come to us with their problems. Before it was not like this. But now they believe that whatever I am saying I have learned from somewhere. They come and show the skin lesion to me, then I just refer them.”* (P11,40’s, Male, Unemployed)

For those with G2D, societal judgment persisted. A couple of young males with hand deformities reported ongoing discrimination and social avoidance in their wider interactions. Others though not physically disabled agreed that social stigma persisted, citing enduring community myths about leprosy and the need for continued awareness efforts.

During photovoice discussions, one participant took this vision further, expressing a wish to connect with persons affected by leprosy in other countries to share stories and experiences.

### 3.3 Process Evaluation of the Storytelling Intervention

#### Enabling Factors

Motivation to join the project stemmed from its **perceived value** for personal growth and its potential for wider societal benefits. Some expressed a desire to share their experiences with purpose, recognising the personal and social value of mastering the skill of storytelling in overcoming stigma. In contrast, an older male with no formal education joined without any particular motives.

**Relational trust** and **psychological safety**, both within the PSG and the broader organisational context, emerged as key enablers of the programme’s effectiveness. Most participants often described a strong sense of connection and inclusivity. This promoted a sense of belonging, which counteracted previous feelings of isolation and made it easier to feel accepted in the group, share openly and stay engaged. Many described the PSG as feeling like home, with its members regarded as family. All respondents highlighted that group activities cultivated unity and solidarity among peers. Outside of the group, being acknowledged and treated as equals, particularly in public platforms when engaging with non–affected persons, was a form of powerful affirmation. **Social recognition** was particularly significant for many of the persons affected, in strengthening identities and amplifying their voices. This was echoed during photovoice discussions:

> *“The important message, which I want to deliver “We are different from others, not less”[…] First start giving respect to yourself if you want to take respect from others. You should identify the inner strength of yourself, hidden strength. First discipline, second focus, third – self-confidence. Your focus should be like an eagle. Self-confidence should be like a lion” (Photovoice discussion)*

#### Challenges

Early engagement was sometimes limited by **psychological barriers**, such as scepticism towards unfamiliar formats and apprehension about emotional disclosure. A few individuals, both men and women, initially feared emotionally vulnerability in group settings and a couple struggled to recall past events, particularly when their diagnosis and treatment had occurred decades earlier.

Prevailing **gender norms** shaped women’s participation and autonomy to engage freely. All required permission from husbands or male relatives to attend and some could only participate if accompanied by a male companion. All female participants voiced concern about mixed-gender settings, regarding how women would express themselves in front of men. In practice, however, they spoke comfortably in this setting and reported finding it useful. Additionally, family attitudes influenced participation, with a few respondents describing how relatives questioned the value of their involvement.

**Logistical constraints**, such as distance, travel costs and time availability, were common challenges for both men and women, particularly for those balancing work and care responsibilities. For example, one man missed the film screening due to his daughter’s sickness, while one woman attended despite her husband’s illness, describing taking multiple buses and spending hours commuting. These practical factors made participating challenging, especially in Karachi’s congested urban setting.

### 3.4 High Impact Components of the Programme

#### Creative narrative tools

Many participants identified creative activities (e.g. poetry, singing) as one of the most meaningful components of the programme. These methods reportedly enabled reflection and expression, leading to emotional release and a sense of relief. They were seen as an engaging way to process and communicate their lived experiences. The ‘River of Life’ exercise was described as particularly powerful, mentioned by several participants in both IDIs and photovoice discussions. In particular, it helped them organise their life stories, express emotions more clearly, and match their body language and voice to what they were feeling.

> *“We learned that if you are sharing your story with someone how it should be, it gives a sequence or an order to your story for better understanding to the listener, so he feels the burden on your heart or he feels very good and better.”* (P12, 60’s, Male, Self-employed)

#### Non-clinical and leisure-based settings

Stepping outside clinical environments was an important aspect of the programme, as reported by over half the respondents. Natural, informal settings (e.g. parks or the beach) reportedly introduced an element of joy that stood in contrast with the hospital’s atmosphere, with respondents often noting they preferred to focus on other aspects of life. This shift was illustrated in the photovoice data, as demonstrated in Figure 4.

**Figure 4.**
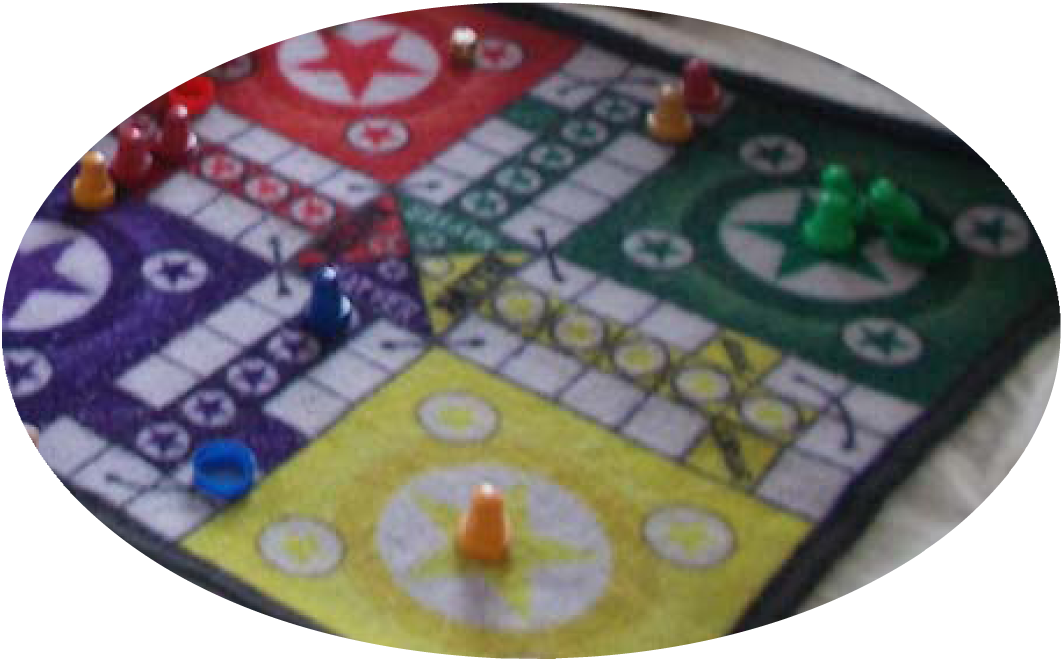
“We learned through games in the project. They helped us forget our sadness and pain, and we had a lot of fun together. When you play games, you can step out of your sorrow or pain - the game distracts you. There should be some entertainment or games so you can forget your pain”.

#### Community exposure and public visibility

Many emphasised community exposure as a key element. Field visits and a film screening were attended by national media outlets, giving participants’ voices greater visibility. High-profile public events, such as World Leprosy Day, increased public awareness about leprosy.

> “*When I was giving a speech, I was feeling like a motivational speaker. I didn’t feel that I am a leprosy patient or I am deformed […] That day I was full of confidence and I presented my speech better because I was feeling no hesitation.”* (P01, 30’s, Male, G2D)

These experiences reportedly elevated participants’ sense of importance and reshaped both how they saw themselves and how others perceived them. Half noted that performing in front of staff also reinforced their confidence and offered recognition beyond their peers.

#### Demographic and social characteristics

Gender shaped experiences of both stigma, empowerment and participation. Women described constraints on attending the project workshop sessions. For men, stigma was tied to threats to their role as providers. All participants in the study were married and for the majority the marital relationship was described as an important source of support in coping with their diagnosis. Visible deformities (G2D) intensified both enacted and anticipated stigma, including public scrutiny and efforts to conceal impairments. Employment context influenced experiences. Those with roles related to leprosy services described some protection within work settings, though broader community stigma persisted.

### 4.5 Participant Recommendations

Here, we present participants’ suggestions to enhance the programme, including amendments and alternative approaches. A critical insight was the need to expand outreach for meaningful, regular contact between affected and non-affected groups, with a strong emphasis on peers leading these initiatives. This included a need to leverage peer diversity by matching roles to skills and language capacities, supporting woman to woman outreach and leveraging a wide range of multi-media channels and settings (e.g., schools). Participants also called for enhancements to programme structure and delivery, including more structured and inclusive recruitment pathways, gender segregated groups, further technical content (e.g., on early treatment of leprosy) and increasing the duration and diversity of activities. Additionally, participants called for the integration of economic empowerment activities and emphasised the importance of prioritising sustainability (e.g., transition to peer leadership and community ownership and resource mobilisation and decentralised hubs). See S8 Appendix for a detailed overview of recommendations and specific activities.

## 4. Discussion

This study sought to explore the impact of a creative storytelling intervention on stigma and empowerment among persons affected by leprosy in Pakistan. Our findings highlight storytelling interventions as a key catalyst for identity transformation and personal empowerment in persons affected by leprosy.

Interpreted through Perkins and Zimmerman’s Psychological Empowerment Framework, shifts from shame and concealment to self-acceptance, confidence and pride reflect enhanced self-efficacy and identity redefinition, the essence of intrapersonal empowerment, while interactional (e.g., communication skills) and behavioural gains (e.g., outreach) were present but secondary (23). Effective stigma reduction requires multi-level, multi-component interventions that engage both stigmatised individuals and those who stigmatise, extending beyond individual change to interpersonal, community, and structural levels to achieve lasting impact (26, 56–58) (59). The symbiotic stigma-empowerment dynamic (Figure 5) created a self-perpetuating cycle of change, underpinned by key psychosocial mechanisms (60, 61).

**Figure 5.**
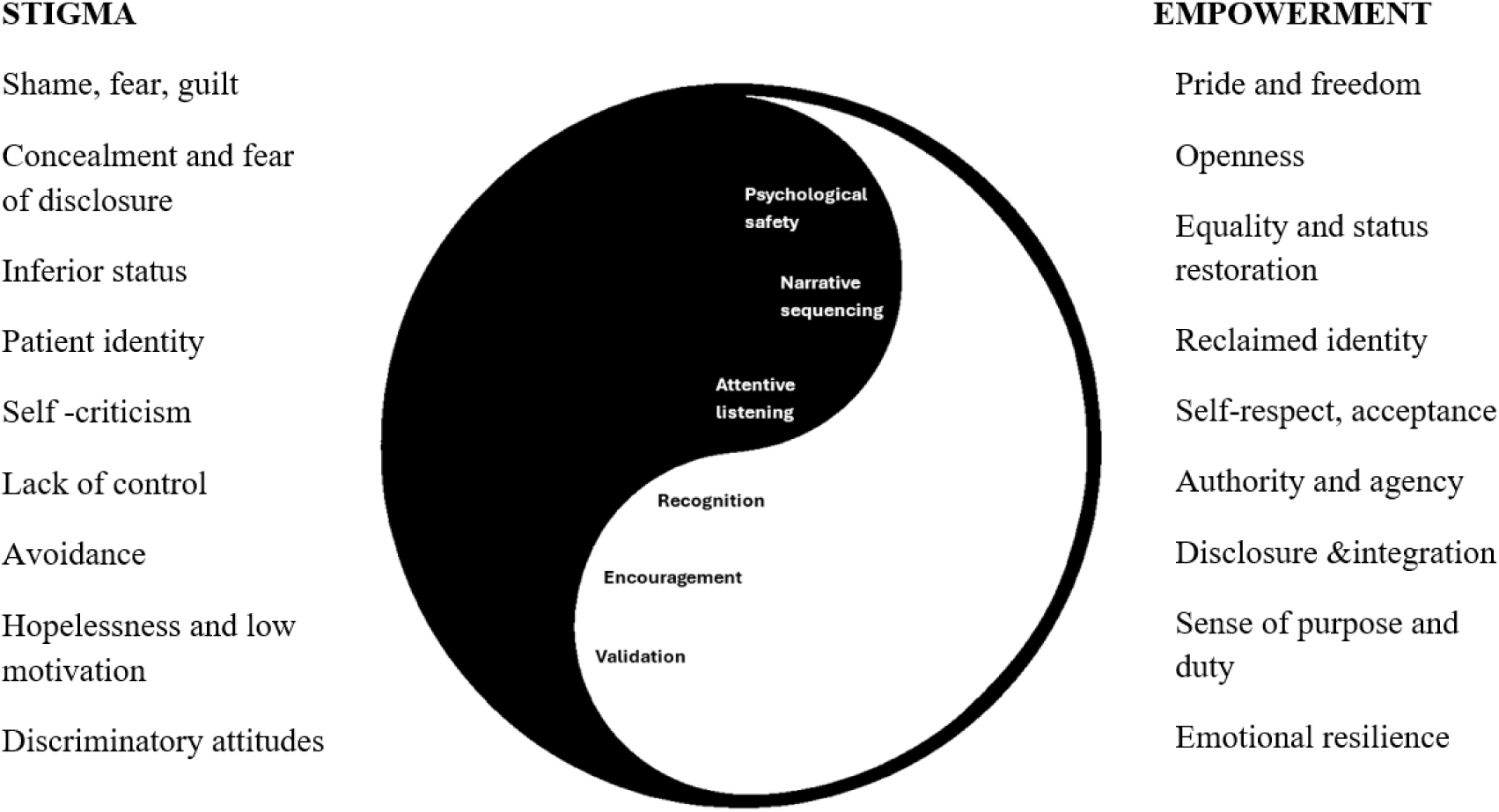
Symbiotic relationship between stigma reduction and enhanced empowerment illustrating the driving factors for personal transformation (Source: Author’s own)

Our findings echo similar mechanisms identified in comparable interventions among persons affected by leprosy. In Indonesia’s SARI project, participatory video enabled identity reframing, from stigmatised patients to capable creators and storytellers, enhancing pride and self-worth (34). Gaining authorship over their narratives, restored their sense of dignity and agency (34). However, unlike SARI, participants in this study had limited control over dissemination, which may have constrained empowerment (43, 62).

The photovoice process appeared to function as a modest extension of the storytelling intervention. By inviting participants to select and interpret images, it enabled them to distil their narratives into visual metaphors that reflected shifts in identity, confidence and anticipated stigma. Group-based discussion and clustering of images and themes enabled collective meaning-making, repositioning participants as co-analysts of their own experiences and providing a safe space for potential advocacy roles in the future. In this way, photovoice both triangulated and deepened the IDI findings on empowerment.

### Peer Support Groups and Institutional Integration are Central

The PSG, embedded in MALC, actively enabled change, bridging the gap between the participants’ current capabilities and potential development (23, 33, 63). This aligns with emphasis on community mobilisation as a pathway to empowerment (64) and highlights the role of social structures in resisting stigma’s marginalising effects (26, 65).Figure 6 illustrates the relational and institutional mechanisms through which PSGs facilitate empowerment.

**Figure 6.**
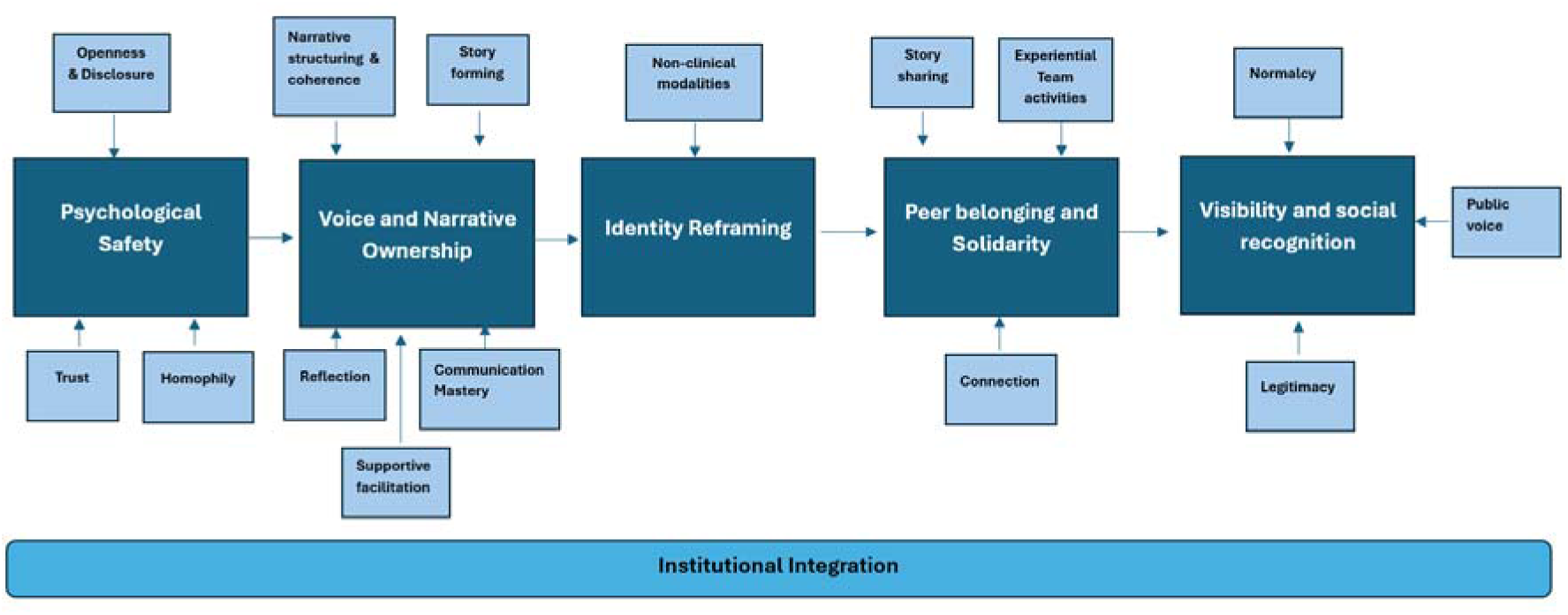
Pathway of mechanisms underlying the storytelling process in relation to the PSG and institution (Source: Author’s own)

MALC’s institutional support (via training and certification) enabled access, conferred legitimacy and positioned participants as credible advocates, mirroring how organisational endorsement strengthens contact conditions (66, 67). Peer support built bonding social capital (68), rooted in empathic listening and shared lived experiences (69). Such interactions reportedly built relational trust, making individuals feel respected and willing to disclose (70, 71). These dynamics transformed individual gains into interactional empowerment, supporting identity reframing, as reported in the literature (72, 73).

Most participants described safe, equal-status peer spaces where encouragement enabled disclosure and public speaking, seen as essential for empowerment (23, 74). This dynamic was evident during the photovoice group discussions, where participants worked together to interpret their images and weave past, present and future into a shared, coherent story of change. Observing these interactions offered insight into how the group functioned collectively, rather than as separate individuals, further illustrating the collaborative foundations of empowerment within the peer group.

Our findings align with evidence on community-based groups (including PSGs) that combine peer support and skills-building to promote empowerment (9, 75, 76). The Hotopf et al. review highlights the role of groups in in positioning persons affected as contributors but notes that participation often stalls at mobilisation and collaboration rather than reaching delegated leadership (77, 78). Our data identifies the same ceiling, with no formalised peer roles, referral pathways or co-planning.

While institutional affiliation, such as MALC’s longstanding reputation, enhanced legitimacy, visibility and participant confidence, it also introduced a subtle dependency. Participants sometimes deferred ownership to the organisation, which constrained their sense of agency and autonomy. This reflects a broader paradox in empowerment practice; institutional support can both enable and limit grassroots empowerment, especially when top-down dynamics overshadow community leadership (79). As such, long-term sustainability requires a shift from institutional facilitation toward community ownership, aligning with the highest levels of participatory engagement (77). Through photovoice discussions, this dynamic became visible; while participants expressed gratitude for MALC’s support, their conversations also included tentative ideas about organising themselves and creating more independent, peer-led spaces.

### Structural Inequalities and Participation: Implications for Equity and Effectiveness

Our findings align with evidence from Pakistan, where patriarchal norms constrain women’s autonomy (37, 80). This highlights the importance of addressing household power dynamics, as they influence healthcare access. Reliance on male decision-makers, as identified in this study, can limit timely and autonomous engagement with interventions (81).

These gendered barriers resonate with reviews documenting triple discrimination and literature showing how cultural norms, mobility constraints and family scrutiny restrict women’s agency in and access to healthcare (37, 74, 82, 83) (84–86). These challenges can further dampen participation and empowerment unless skin NTD programmes adapt (27, 87). However, where supportive conditions existed in the programme, such as staff and peer encouragement, as reported by most women, participants described greater gains, including reduced shyness and increased feelings of ‘boldness’.

Visible impairments (G2D), particularly among males, remain a salient factor in shaping exposure to enacted stigma, through their association with heightened contagion fears (88). This pattern persists regardless of social or professional standing, aligning with existing evidence (17, 89). Early treatment and self-care in leprosy are therefore critical, not only to improve health outcomes but also reduce the risk of disability, which in itself exacerbates stigma (90).

Socioeconomic status acted as both a driver and an outcome of stigma, supporting existing literature (27, 78). Discrimination and job loss lead to poverty, which restricts access to healthcare, reinforcing a cycle of exclusion (88, 91, 92). Although only a couple of participants raised the need for livelihood opportunities, their accounts highlight that economic inclusion is central to achieving equity for persons affected by leprosy. Unemployed participants faced financial barriers (e.g., transport costs), while those with jobs had resources but less time, echoing similar challenges in the literature, underscoring the need for flexible, supported participation (83, 93).

The demand for livelihood opportunities echoes empirical findings. In Indonesia, combining microcredit with social support reduced stigma and improved economic security and quality of life (94, 95), though the study’s small sample limits generalisability. The MALC PSG could serve as a vehicle for such initiatives (78). This approach is reinforced by the WHO Global Leprosy Strategy 2021–2030, which positions social and economic rehabilitation alongside medical care as essential to achieving zero stigma and full social participation (11).

### Addressing Persistent Community-level Stigma: The Role of Contact, Reach and Repetition

At the community level, gains were selective, limited to MALC-linked settings and informal dialogue, while public misconceptions and enacted stigma persisted, especially outside of the ‘safe’ organisational setting. This aligns with evidence of widespread community stigma and social avoidance in Pakistan (22, 37, 38, 96).

As Willis et al. note in their systematic review, most leprosy stigma interventions target affected persons rather than wider communities (97). In the context of our study, this risks confining change to organisational settings without shifting broader community norms, thereby revealing the limits of isolated interventions against entrenched social practices. Limited reach may also reflect the nature of contact opportunities within the programme. While several participants described equal-status engagement and institutional endorsement, conditions aligned with Allport’s theory for positive intergroup contact (66), these interactions were episodic, institution-bound and reached a narrow audience.

Contact, both direct and indirect, is among the most effective stigma-reduction strategies, but its impact is greatest when frequent, sustained, tailored to the audience and extended beyond institutional confines (15, 27, 57, 98). Corrigan et al. argue the importance of targeting specific groups (e.g. HCPs and students), through direct contact led by people with lived experience, coupled with follow up actions (99). This echoes most participants’ call for increased community exposure and dialogue, particularly in schools and high-prevalence communities.

Although the storytelling intervention incorporated IEC components during camp visits, these were largely standalone. Evidence shows that while IEC can improve knowledge, it rarely leads to sustained attitude or behaviour change on its own (25, 100). Instead, integrated approaches combining IEC with structured contact are more effective (9).

### Evidence-based Recommendations

Below, we present a set of evidence-informed recommendations for an integrated approach to stigma reduction and empowerment, based on four intersecting components: storytelling, community-based groups, IEC strategies, and contact-based elements, with persons affected at its core (Table 2). Future stigma-reduction programmes should prioritise peer-led storytelling across the full project cycle, combining sustained community contact with wider dissemination to extend impact beyond organisational settings. Embedding interventions within institutional and health system structures can enhance legitimacy, sustainability, and scale, particularly when peer leadership is formalised through recognised roles and pathways. To translate empowerment into durable inclusion, programmes should integrate capacity-building with economic opportunities via local partnerships. Gender- and disability-responsive design is essential, ensuring women and persons with G2D are positioned in visible leadership roles, household barriers are addressed, and participation models remain flexible and context-sensitive. Such actions are critical as we continue striving towards realising the Road Map for NTD 2021-2030 targets (101), including the implementation of the Essential care package to address mental health and stigma for persons with NTDs (102).

**Table 2:** Recommendations.

| **Recommendation** | **Specific Activities** |
| --- | --- |
| Enhance peer-led storytelling and establish regular community contact | • Adopt asset-based, peer-driven models that embed participant involvement from design through evaluation (33, 103-106)<br>• Combine structured preparatory IEC with recurring contact opportunities across community and institutional settings (33, 95, 98, 107)<br>• Expand digital dissemination of creative outputs led by peers (31). |
| Integrate programmes into institutional structures | • Embed stigma-reduction interventions within existing health systems, NTD services, and local organisations, ensuring representation of persons affected in advisory, technical, |
|  | and guideline-setting structures (11, 40, 98, 102–104).<br>• Formalise peer networks and leadership through structured training, defined peer roles, accreditation, and supervision, aligned with established models and Pakistan’s Roadmap towards Zero Leprosy (8, 61, 88, 105). |
| Strengthen capacity and economic inclusion through local partnerships | • Strengthen pathways to economic inclusion by linking peer-supported empowerment initiatives with employment and livelihood opportunities through local partnerships (78, 108) (11, 90, 94, 108, 109). |
| Integrate gender and disability inclusion into programme design | • Ensure female-led peer recruitment with visible roles, using flexible participation models and family engagement to address household and gender-based barriers (110)<br>• Include persons affected with G2D in leadership and public facing roles to disrupt stigma cues, confront societal discomfort and challenges perceptions that equate deformity with danger or incapacity (15, 111, 112). |

### Study strengths and limitations

This study makes a novel contribution to the limited evidence on the impact of storytelling interventions among persons affected by leprosy. Findings and recommendations have the potential to support equitable, effective storytelling interventions in other skin NTDs. The use of multiple participatory methods centred the lived experience of persons affected, supporting the credibility of our findings. However, there are some limitations to consider.

Firstly, we used a relatively small sample, though saturation was reached, demonstrating the credibility of our findings, despite the small sample (113). Perceived empowerment gains or understated ongoing stigma could be inflated since no participant could be recruited that did not complete the intervention. Gender representation was uneven, which limited insight into gendered dynamics (84). However, the analysis attended to how gender shaped experiences ensuring women’s perspectives informed interpretation (114). Moreover, newly diagnosed individuals were also under-represented, in the sample and within the intervention itself due to declining incidence in the region, which limits our understanding of intervention efficacy across the diagnostic timeline (115–117).

Furthermore, whilst conducting interviews at MALC headquarters, as well as involving programme staff, offered a familiar setting, this may have signalled institutional authority, potentially heightening social desirability bias and prompting responses shaped by organisational expectations, however, mitigated by anonymisation (118, 119). Language barriers and translation added complexity, with interpreting between Urdu and English sometimes introducing semantic discrepancies (120). To mitigate this, interviews included real-time summaries to confirm accuracy and reporting used thick description to preserve context and transferability (121).

For future research, controlled or randomised designs with matched comparison groups would strengthen causal inference and provide a more rigorous assessment of impact and feasibility. Moreover, an enlarged scope would offer further insights, including diversifying and expanding the group of participants, as well as assessing long-term impacts of such interventions.

## Conclusion

This study provides evidence that a participatory storytelling intervention in a low-endemic context in Pakistan effectively shifted self-stigma and intrapersonal empowerment. Moreover, the modality of the PSG and the institutional embedding shapes feasibility and uptake. An integrated stigma-empowerment mode was generated from the study, which lays a pathway for sustainable progress, requiring a sequenced, multi-level strategy, which can be applied in future person-centred research, including the implementation of the Essential Care Package for mental health and stigma. The findings implicate and formulate the call for specific attention to women’s participation and disability inclusion to ensure an equitable process and outcome. Moreover, intervention spaces should be expanded through regular, targeted contact with defined audiences, advanced by accredited community leaders, as well as strengthened evaluation. Although transferability is plausible, application should be cautious and contingent on adaptation and testing in other settings.

## Data Availability

All relevant data are within the manuscript and its supporting information files.

## Acknowledgements

To all the participants, thank you for the privilege of sharing your stories and experiences with us. Your voices are the foundation of this work. We also give thanks to Dr Nara Tagiyeva-Milne, Mia Lindfield and Alison Derbyshire at LSTM and to Mr Abdul Salam who was integral during the field work.

